# Endothelial cell-activating antibodies in COVID-19

**DOI:** 10.1101/2021.01.18.21250041

**Authors:** Hui Shi, Yu Zuo, Sherwin Navaz, Alyssa Harbaugh, Claire K. Hoy, Alex A. Gandhi, Gautam Sule, Srilakshmi Yalavarthi, Kelsey Gockman, Jacqueline A. Madison, Jintao Wang, Melanie Zuo, Yue Shi, Michael D. Maile, Jason S. Knight, Yogendra Kanthi

## Abstract

**Objective:** While endothelial dysfunction has been implicated in the widespread thrombo-inflammatory complications of coronavirus disease-19 (**COVID-19**), the upstream mediators of endotheliopathy remain for the most part cryptic. Our aim was to identify circulating factors contributing to endothelial cell activation and dysfunction in COVID-19.

**Methods:** Human endothelial cells were cultured in the presence of serum or plasma from 244 patients hospitalized with COVID-19 and plasma from 100 patients with non-COVID sepsis. Cell adhesion molecules (E-selectin, VCAM-1, and ICAM-1) were quantified by in-cell ELISA.

**Results:** Serum and plasma from patients with COVID-19 increased surface expression of cell adhesion molecules. Furthermore, levels of soluble ICAM-1 and E-selectin were elevated in patient serum and tracked with disease severity. The presence of circulating antiphospholipid antibodies was a strong marker of the ability of COVID-19 serum to activate endothelium. Depletion of total IgG from antiphospholipid antibody-positive serum markedly restrained upregulation of cell adhesion molecules. Conversely, supplementation of control serum with patient IgG was sufficient to trigger endothelial activation.

**Conclusion:** These data are the first to suggest that some patients with COVID-19 have potentially diverse antibodies that drive endotheliopathy, adding important context regarding thrombo-inflammatory effects of autoantibodies in severe COVID-19.

## INTRODUCTION

There are several likely synergistic mechanisms by which SARS-CoV-2 infection may result in COVID-19-associated coagulopathy including cytokine release that activates leukocytes, endothelium, and platelets; direct activation of various cells by viral infection; and high levels of intravascular neutrophil extracellular traps (**NETs**) (1). The latter are inflammatory cell remnants that amplify thrombosis (2). COVID-19-associated coagulopathy may manifest with thrombosis in venous, arterial, and microvascular circuits. The incidence of venous thromboembolism is particularly notable in severe COVID-19 (10% to 35%) with autopsy series suggesting that as many as 60% of those who succumb to COVID-19 are impacted (3).

Recently, there have been a number of descriptions of what appears to be *de novo* autoantibody formation in individuals with severe COVID-19. One example replicated by multiple groups is the detection of antibodies reminiscent of the antiphospholipid antibodies (**aPL**) that mediate antiphospholipid syndrome (**APS**) in the general population. In APS, patients form durable autoantibodies to phospholipids and phospholipid-binding proteins such as prothrombin and beta-2-glycoprotein I (**β_2_GPI**). These autoantibodies then engage cell surfaces, where they activate endothelial cells, platelets, and neutrophils and thereby tip the blood:vessel wall interface toward thrombosis. While viral infections have long been known to trigger transient aPL (4), mechanisms by which these potentially short-lived antibodies may be pathogenic have not been deeply characterized. To this end, our group recently found that IgG fractions from patients with COVID-19 were enriched for aPL and potentiated thrombosis when injected into mice (5). Intriguingly, the circulating B cell compartment in COVID-19 appears similar to the autoimmune disease lupus, whereby naïve B cells rapidly take an extrafollicular route to becoming antibody-producing cells (6), and in doing so bypass the normal tolerance checkpoints against autoimmunity provided by the germinal center.

Here, we were initially interested in the extent to which circulating NET remnants might be an important activator of endothelial cells. We then also necessarily turned our attention to aPL as potential markers of COVID-19 serum and polyclonal IgG fractions with strong endothelial cell activating-potential.

## MATERIALS AND METHODS

Detailed methods are available in the Supplement. This study complied with all relevant ethical regulations and was approved by the University of Michigan IRB (HUM00179409 and HUM00131596).

### Serum and plasma samples from patients with COVID-19

All 118 patients in the primary cohort and 126 patients in the expansion cohort had a confirmed COVID-19 diagnosis based on FDA-approved RNA testing.

### Cell culture

Human umbilical vein endothelial cells (**HUVEC**) were cultured in Endothelial Cell Growth Basal Medium-2 (CC-3156, Lonza) supplemented with EGM-2 MV SingleQuots Kit (CC-4147, Lonza).

### Purification of human IgG

IgG was purified as we have described previously (5).

### In-cell ELISA and neutrophil adhesion

In-cell ELISA and neutrophil adhesion were by protocols that we have described previously (7).

### Quantification of antibodies and biomarkers

aPL were quantified using Quanta Lite® kits (Inova Diagnostics Inc.). Soluble E-selectin and ICAM-1 were quantified by ELISA (DY724 and DY720, R&D Systems). Myeloperoxidase-DNA complexes were quantified as has been previously described (8). Cell-free DNA was quantified using the Quant-iT PicoGreen dsDNA Assay Kit (Invitrogen, P11496). Citrullinated histone H3 was quantified by ELISA (Cayman, 501620). Calprotectin was quantified by ELISA (DY8226, R&D Systems).

## RESULTS

### Activation of HUVEC by COVID-19 serum

Serum samples were collected from 118 patients hospitalized with COVID-19 at an academic hospital. The average age of this cohort was 62, while 47% were female and 42% were Black/African American (**Supplemental Table 1**); 35% were receiving mechanical ventilation. Serum from COVID-19 patients were added to early-passage HUVEC, and the expression of cell adhesion molecules was determined after 6 hours via a custom in-cell ELISA compatible with the Biosafety Level-3 facility per institutional guidelines (**Figure 1A**). As compared with serum samples from 38 healthy controls, the COVID-19 samples triggered an activated endothelial cell phenotype as evidenced by increased surface expression of the cell adhesion molecules E-selectin (**Figure 1B, Supplemental Figure 1A**), VCAM-1 (**Figure 1C, Supplemental Figure 1B**), and ICAM-1 (**Figure 1D, Supplemental Figure 1C**). Relative activation as compared with unstimulated cells was similar to that observed with 20 ng/ml TNF-α stimulation, which was included in all experiments as a positive control (**Supplemental Figure 2**).

**Figure 1:**
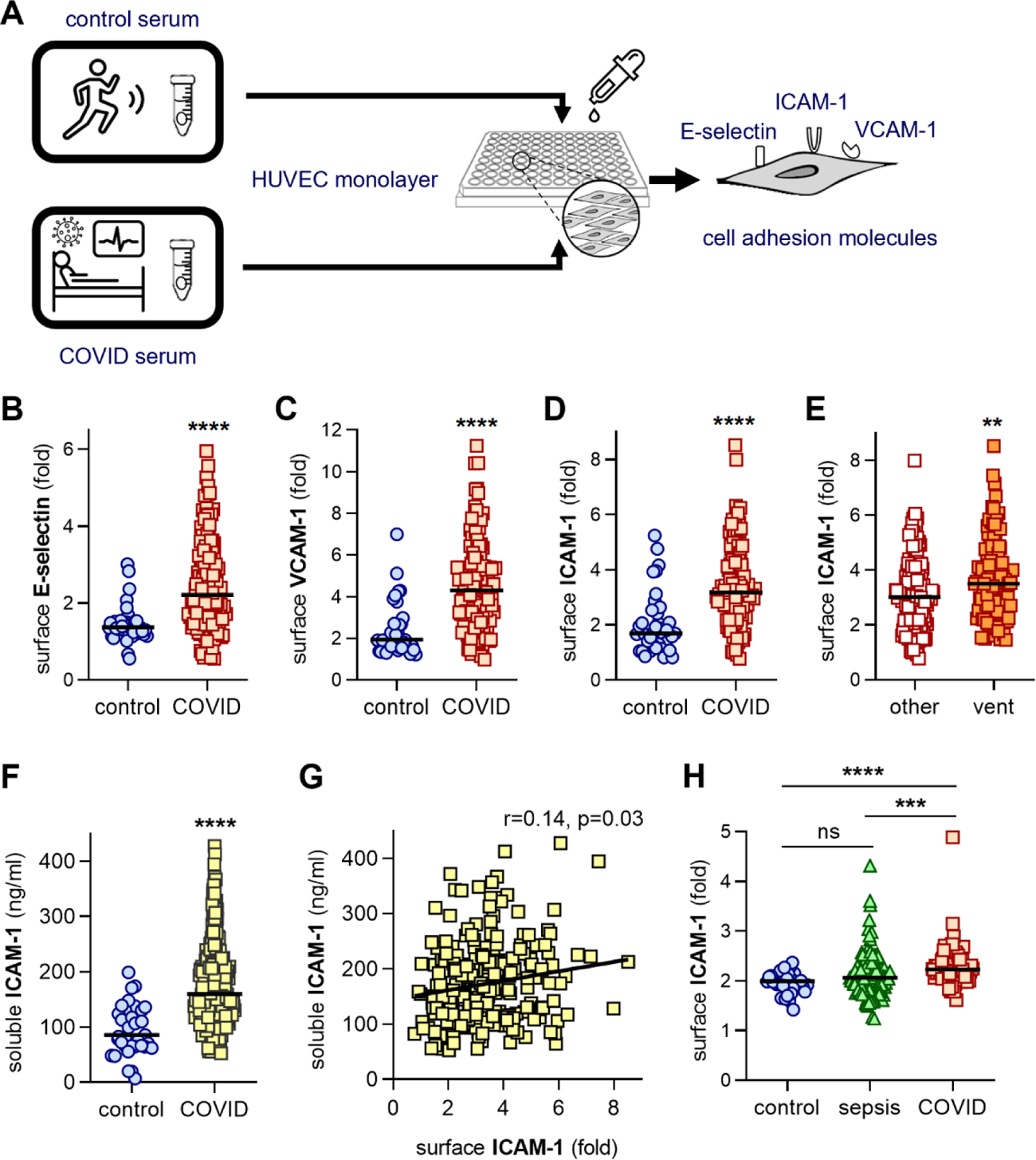
Activation of human umbilical vein endothelial cells (HUVEC) by control or COVID-19 serum. **A**, Schematic workflow for in-cell ELISA. HUVEC were cultured for 6 hours with serum from either healthy controls (collected pre-pandemic) (n=38) or patients hospitalized with COVID-19 (n=118). Cells were then fixed, and surface expression of E-selectin (**B**), VCAM-1 (**C**), or ICAM-1 (**D**) was quantified. Median values are indicated by horizontal lines. Groups were analyzed by Mann-Whitney test; ****p<0.0001. **E,** Beyond the 118 COVID-19 patients tested in panel D, surface expression of ICAM-1 was tested in the context of an additional 126 unique patient samples. Patients requiring mechanical ventilation (n=101) are compared to hospitalized patients who were not mechanically ventilated (n=143); **p<0.01 by Mann Whitney test. **F,** Serum from healthy controls (n=37) and COVID-19 patients (n=232) were assessed for soluble ICAM-1. COVID-19 samples were compared to controls by Mann-Whitney test; ****p<0.0001. **G,** Soluble ICAM-1 (n=232) was compared to HUVEC ICAM-1 expression as presented in panel E. Correlation was determined by Spearman’s method. **H,** HUVEC were cultured for 6 hours with plasma from healthy controls (n=36), intensive care unit patients with non-COVID sepsis (n=100), or patients hospitalized with COVID-19 (n=72). Cells were then fixed and surface expression of ICAM-1 was quantified. Groups were analyzed by Kruskal-Wallis test with correction for multiple comparisons by Dunn’s method; ***p<0.001, ****p<0.0001, and ns=not significant.

We were able to obtain sufficient serum from an additional 126 hospitalized COVID-19 patients to expand the ICAM-1 testing. These patients had a similar profile to the original 118 patients as shown in **Supplemental Table 1**, and also in terms of upregulating surface ICAM-1 (**Supplemental Figure 3**). Considering all 244 patients together, serum from patients requiring mechanical ventilation upregulated ICAM-1 more strongly that did serum from patients who were hospitalized but not requiring mechanical ventilation (p<0.01, **Figure 1E**). We also measured levels of soluble ICAM-1 and E-selectin in patient serum. We found significantly higher levels of both soluble ICAM-1 and E-selectin in COVID-19 serum as compared with healthy controls (**Figure 1F** and **Supplemental Figure 4**). Soluble ICAM-1 demonstrated a positive correlation with HUVEC ICAM-1 expression (**Figure 1G**), as well as with clinical parameters that track with COVID-19 severity including C-reactive protein (r=0.27, p=0.0002), D-dimer (r=0.29, p=0.0001), and oxygenation efficiency (r=-0.37, p<0.0001, **Supplemental Figure 5**). Soluble E-selectin demonstrated similar correlations with all of the above (**Supplemental Figure 4**).

To determine the extent to which this phenomenon might extend to critically ill patients without COVID-19, we matched 100 patients with sepsis and admitted to the intensive care (plasma obtained in the pre-COVID era) to our COVID-19 cohort (n=72 plasma samples available) by age and comorbidities (**Supplemental Table 2**). More patients in the sepsis cohort required mechanical ventilation (65% versus 46%). We then stimulated HUVEC with plasma and determined the surface expression of ICAM-1. While a subset of the sepsis plasma elicited high expression of surface ICAM-1, the group as a whole was not significantly different from the controls (**Figure 1H**). Meanwhile, COVID-19 plasma demonstrated more activation than either control or sepsis plasma (**Figure 1H**). Taken together, these data indicate that COVID-19 serum and plasma contain factors capable of activating endothelial cells.

### Association of NET remnants and other biomarkers with the endothelial cell-activating potential of COVID-19 serum

Given that NETs are known activators of endothelial cells, we asked whether NET remnants might predict the ability of a particular COVID-19 serum sample to upregulate surface E-selectin, VCAM-1, and ICAM-1. Specifically, we measured NETs in COVID-19 serum (n=118) via quantification of cell-free DNA, myeloperoxidase (MPO)-DNA complexes, and citrullinated histone H3 (**Cit-H3**) (**Table 1**). Serum MPO-DNA complexes demonstrated modest correlations with surface expression of both VCAM-1 (r=0.26, p<0.01) and ICAM-1 (r=0.28, p<0.01). Cell-free DNA and Cit-H3 both correlated with VCAM-1 only (r=0.24, p<0.05 and r=0.22, p<0.05, respectively) (**Table 1**). Beyond these relatively specific markers of NETs, we also sought correlations with more general markers of thrombo-inflammation including C-reactive protein, D-dimer, and calprotectin, the latter having been previously shown to be an early predictor of respiratory failure in COVID. Of the three, only calprotectin demonstrated positive correlations with expression of all three endothelial cell surface markers (r=0.19-0.20, p<0.05), while D-dimer correlated with ICAM-1 only (r=0.26, p<0.05) (**Table 1**). Taken together, these data demonstrate modest correlations between NETs/thrombo-inflammation and the ability of COVID-19 serum to activate endothelial cells.

**Table 1:** Correlation of HUVEC cell adhesion molecules with antiphospholipid antibodies in COVID-19 patients

|  |  |  |  |  |  |  |
| --- | --- | --- | --- | --- | --- | --- |
|  | E-selectin |  | VCAM-1 |  | ICAM-1 |  |
| Spearman | r | p | r | p | r | p |
| Antiphospholipid antibodies |  |  |  |  |  |  |
| anticardiolipin IgG | 0.446 | **** | 0.421 | **** | 0.346 | *** |
| anticardiolipin IgM | 0.369 | **** | 0.252 | ** | 0.357 | **** |
| anti-β<sub>2</sub>GPI IgG | 0.156 | ns | 0.213 | * | 0.076 | ns |
| anti-β<sub>2</sub>GPI IgM | 0.009 | ns | 0.047 | ns | 0.150 | ns |
| anti-PS/PT IgG | 0.432 | **** | 0.252 | ** | 0.299 | *** |
| anti-PS/PT IgM | 0.254 | ** | 0.115 | ns | 0.276 | ** |
| NET remnants |  |  |  |  |  |  |
| cell-free DNA | 0.073 | ns | 0.237 | * | 0.135 | ns |
| MPO-DNA | 0.156 | ns | 0.256 | ** | 0.277 | ** |
| Cit-H3 | 0.079 | ns | 0.224 | * | 0.076 | ns |
| Other biomarkers |  |  |  |  |  |  |
| C-reactive protein | -0.008 | ns | 0.173 | ns | 0.003 | ns |
| D-dimer | 0.175 | ns | 0.133 | ns | 0.258 | * |
| calprotectin | 0.206 | * | 0.203 | * | 0.197 | * |
| ns=not significant; NET=neutrophil extracellular trap; MPO=myeloperoxidase; Cit-H3=citrullinated histone H3; *p<0.05, **p<0.01, ***p<0.001, and ****p<0.0001 |  |  |  |  |  |  |

### Association of aPL with the endothelial cell-activating potential of COVID-19 serum

We next reasoned that COVID-19-associated autoreactive antibodies might activate endothelial cells. In pursuit of such antibody fractions, we focused on the IgG and IgM isotypes of three types of aPL: anticardiolipin, anti-β_2_GPI, and anti-phosphatidylserine/prothrombin (**anti-PS/PT**). As is detailed in Supplemental Table 3, 45% of subjects were positive for at least one antibody based on the manufacturer’s cutoff and 25% were positive with a more stringent cutoff of ≥40 units. The vast majority of positive results were either anticardiolipin (IgG=3% and IgM=25% of the cohort, respectively) or anti-PS/PT (IgG=24% and IgM=15% of the cohort, respectively). It should also be noted that there were strong correlations amongst many of the antibodies, for example anticardiolipin IgG and anti-PS/PT IgG (r=0.51, p<0.0001) and anticardiolipin IgM and anti-PS/PT IgM (r=0.53, p<0.0001); all correlations are detailed in **Supplemental Table 4**. Furthermore, aPL levels tracked to some extent with clinical biomarkers, especially levels of circulating NETs (**Supplemental Table 5**). None of the aPL were detected at appreciable levels in the healthy control serum (2 positive tests) and plasma (1 positive test) used in this study (Supplemental Tables 6 and 7).

Interestingly, we detected strong correlations between anticardiolipin and anti-PS/PT antibodies and the three markers of endothelial cell activation (E-selectin, VCAM-1, and ICAM-1) (**Table 1**). The only correlation that was not statistically significant was between anti-PS/PT IgM and VCAM-1 (**Table 1**). We additionally performed logistic regression analysis after setting a positive/negative threshold for cell adhesion molecule upreguation that was 2 standard deviations above the control mean (**Supplemental Table 8**); anticardiolipin IgG levels were significantly higher in E-selectin- and VCAM-1-positive patients (p=0.001 and p=0.003, respectively), while anti-PS/PT IgG levels were significantly higher in ICAM-1-positive patients (p=0.018). We also measured the same aPL in the aforementioned sepsis cohort (Supplemental Table 9). Of the 100 patients, 29 were positive for any aPL with 23 positive at the more stringent cutoff of ≥40 units. In contrast to the COVID-19 cohort, the vast majority of positive tests were for anti-PS/PT IgM, the levels of which demonstrated a correlation with the ability of plasma to upregulate ICAM-1 (r=0.29, p<0.01, **Supplemental Table 10**). In summary, we detected correlations between various aPL and endothelial cell activation in COVID-19, and to some extent in sepsis.

### Increased neutrophil adhesion upon serum-mediated HUVEC activation

To determine whether upregulation of E-selectin, VCAM-1, and ICAM-1 is associated with increased adhesive functions of HUVEC, we performed a neutrophil adhesion assay. We pooled serum from three COVID-19 patients with positive anticardiolipin IgG (mean=29 GPL) and separately five COVID-19 patients with positive anti-PS/PT IgG (mean=54 units) activity. We then stimulated HUVEC with the pooled serum. Compared with control serum, both COVID samples significantly increased adhesion of neutrophils to endothelial cells (**Figure 2A**).

**Figure 2:**
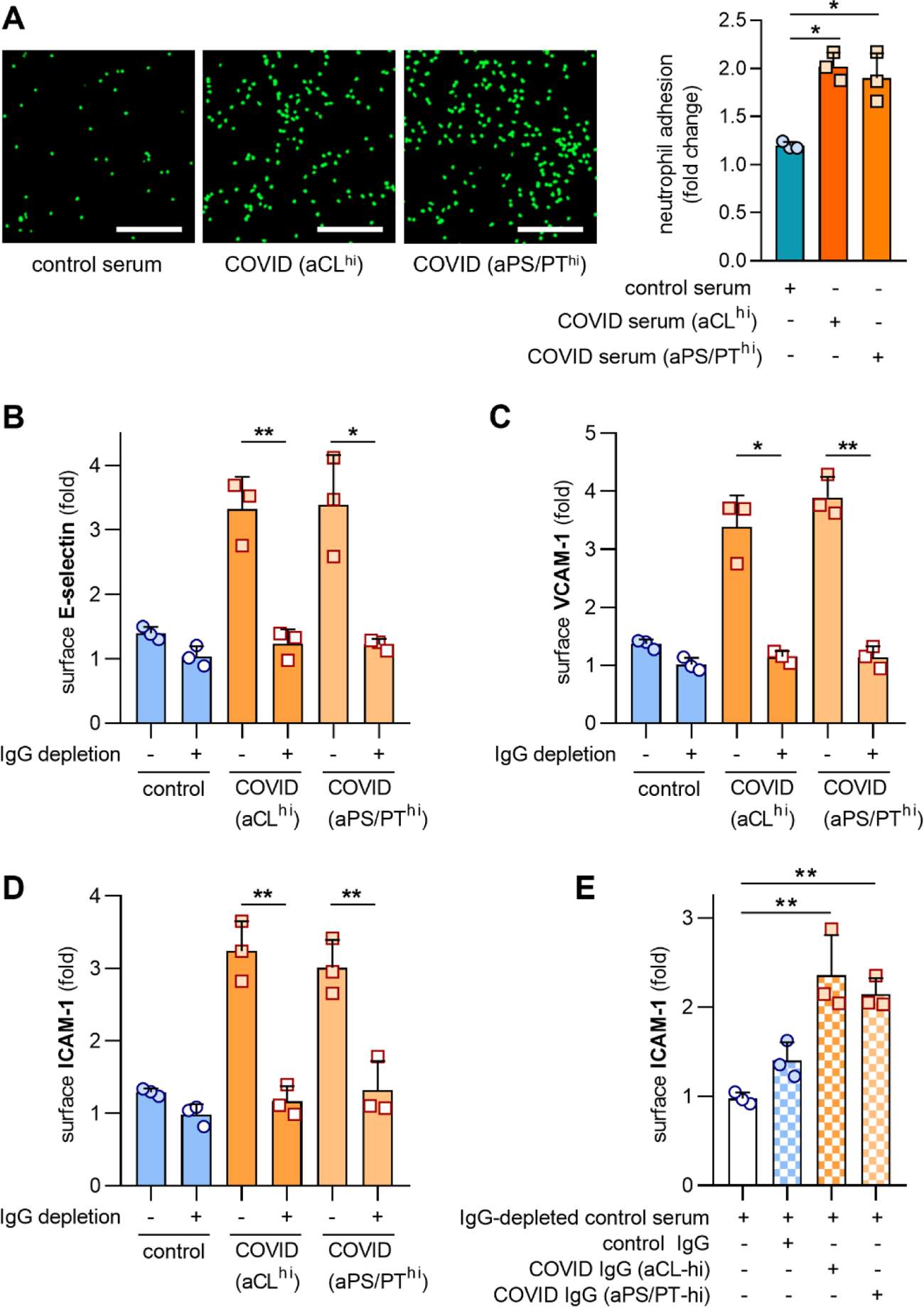
Depletion of IgG from sera with high levels of anticardiolipin (aCL) and anti-phosphatidlyserine/prothrombin (aPS/PT) antibodies alleviates HUVEC activation. A, Serum was pooled from 3 patients with positive aCL IgG, 5 patients with positive aPS/PT IgG, or 3 healthy donors. HUVEC monolayers were treated with 2.5% COVID or control serum for 4 hours. Calcein-AM-labeled neutrophils were then added as described in Methods. Scale bar=200 microns. Mean ± standard deviation is presented for n=3 independent experiments; *p <0.05 by one-way ANOVA corrected by Dunnett’s test. B-D, IgG was depleted from each of the aforementioned pools. Activation of HUVEC was determined after culture for 6 hours as defined by surface expression of E-selectin (B), VCAM-1 (C), or ICAM-1 (D). The experiment was repeated on 3 different days, and bars represent mean and standard deviation. Groups were compared by 2-sided paired t-test; *p<0.05 and **p<0.01. E, IgG (100 μg/ml) was purified from the pooled samples referenced in A-C, and then spiked into control serum that had been depleted of IgG. Activation of HUVEC was determined after culture for 6 hours as defined by surface expression of ICAM-1. Groups were compared by one-way ANOVA with correction for multiple comparisons by Tukey’s test; **p<0.01.

### Depletion of IgG alleviates HUVEC activation

We next sought to determine the extent to which total IgG fractions from COVID-19 patients could directly activate HUVEC. To this end, we subjected the aforementioned pooled serum (anticardiolipin IgG-positive and anti-PS/PT IgG-positive) to either mock or IgG depletion. IgG depletion did not result in significant removal of other serum proteins of note such as calprotectin (**Supplemental Figure 6**). As compared with mock depletion, total IgG depletion completely abrogated the ability of both pooled samples to upregulate endothelial cell E-selectin (**Figure 2B**), VCAM-1 (**Figure 2C**) and ICAM-1 (**Figure 2D**). We next purified a small quantity of total IgG from each pool and added it to control serum that had been depleted of its own IgG. The COVID-19 IgG-supplemented control serum now demonstrated an ability to increase expression of surface ICAM-1 (**Figure 2E**). In contrast, IgG purified from COVID-19 serum that tested negative for both anticardiolipin and anti-PS/PT IgG did not increase expression of ICAM-1 (**Supplemental Figure 7**). Taken together, these data indicate that the presence of aPL correlates with COVID-19 IgG samples that can activate endothelial cells.

## DISCUSSION

Here, we report that serum from COVID-19 patients activate cultured endothelial cells to express surface adhesion molecules integral to inflammation and thrombosis, namely ICAM-1, E-selectin, and VCAM-1. Furthermore, we found that for at least a subset of serum samples from patients with COVID-19, this activation could be mitigated by depleting total IgG. The role of aPL in activating endothelial cells has been demonstrated both *in vitro* and *in vivo* (9). For example, IgG fractions from APS patients have long been known to activate HUVEC, as reflected by increased monocyte adherence and expression of adhesion molecules. It is intriguing that while most characterization of endothelium in the APS field has focused on activation of the endothelium by anti-β_2_GPI antibodies (10), these were only rarely detected in our cohort. Interesting recent work demonstrates the ability of phospholipid-binding, “cofactor-independent” antibodies to also activate endothelium in APS (11). Of course, it should be noted that all experiments performed here were with total IgG fractions and not affinity-purified aPL. Therefore, aPL may mark antibody profiles in severe illness, quite possibly polyclonal, that activate the endothelium and steer the normally quiescent blood:vessel interface toward inflammation and coagulation. In addition to our findings (5), several other groups (12) have also provided evidence for the *de novo* formation of pathogenic autoantibodies in COVID-19. For example, an interesting study used a high-throughput autoantibody discovery technique to screen a cohort of COVID-19 patients for autoantibodies against 2,770 extracellular and secreted proteins (13). The authors found a tendency for autoantibodies to be directed against immunomodulatory proteins including cytokines, chemokines, complement components, and cell surface proteins. This is further bolstered by our data suggesting correlations amongst aPL species in serum from patients with COVID-19.

Beyond COVID-19, we were intrigued to find that approximately one-quarter of sepsis patients had at least one positive aPL test, mostly anti-PS/PT IgM. Given that anti-PS/PT IgM levels tracked with the ability of plasma to activate endothelial cells, we posit that a similar autoreactive antibody-mediated, endothelial cell-activating mechanism may occur in some patients with sepsis, a state in which infection and the host immune response conspire to perturb the vasculature (14). Of particular note, infections causing critical illness have long been known to be potential triggers of autoantibodies, and in particular aPL (4, 15). Although infection-associated aPL have typically been described as transient (16), a recent systematic review found that one-third of individuals found to have positive aPL in the setting of a virus-associated thrombotic event continue to have durably-positive aPL for at least several months (15). Based on our data, studies with long-term clinical and serological follow-up of patients may be necessary to better define the natural history of COVID-19 and non-COVID sepsis.

There are several potential clinical implications of our findings. A consideration that warrants further investigation is whether patients with severe COVID-19 should be screened for aPL to evaluate their risk of thrombosis and progression to respiratory failure, and whether patients with high aPL titers might benefit from treatments used in traditional cases of severe APS such as therapeutic anticoagulation, complement inhibition, and plasmapheresis. At the same time, determining the extent to which aPL are direct mediators of the endothelial cell phenotypes observed here, or perhaps highlight polyclonal antibody fractions likely to activate endothelial cells, is an important question deserving of future research. This study has additional limitations including a lack of a direct readout of macrovascular thrombosis available in other well-conducted studies (17) given aggressive anticoagulation used at our center early in the pandemic, and an as yet incomplete understanding of mechanisms by which aPL-associated IgG fractions activate endothelial cells. However, given the urgency of COVID-19 research, we believe these issues are counterbalanced by the relatively large sample size and the heretofore unknown discovery of endothelial cell-activating antibody profiles in some COVID-19 serum.

Indeed, these data also put the diffuse organ involvement of COVID-19 into context, where a non-specific humoral response to the illness may disrupt the normally quiescent endothelium and potentiate vascular inflammation. As we await definitive solutions to the pandemic, these findings add important context to the complex interplay between SARS-CoV-2 infection, the human immune system, and vascular immunobiology.

## Data Availability

Upon publication in a peer-reviewed journal, data will be made available by the corresponding authors upon request.

## ACKNOWLEDGMENTS

The authors are indebted to all the individuals with COVID-19 and sepsis who participated in this study, and to the frontline workers during the pandemic.

## SUPPLEMENTARY INFORMATION

## MATERIALS AND METHODS

### Serum and plasma samples from patients with COVID-19 and sepsis

Blood was collected into either serum separator tubes containing clot activator and serum separator gel or EDTA tubes by a trained hospital phlebotomist. After completion of testing ordered by the clinician, the samples were released to the research laboratory. Serum and plasma samples were immediately divided into small aliquots and stored at −80°C until the time of testing. All 118 patients in the primary cohort and 126 patients in the expansion cohort had a confirmed COVID-19 diagnosis based on U.S. Food and Drug Administration (FDA)-approved RNA testing. All COVID-19 aspects of the study complied with all relevant ethical regulations and was approved by the University of Michigan Institutional Review Board (HUM00179409). For sepsis, consent was obtained from either the patient or their legally authorized representative (HUM00131596). Blood was collected through an existing catheter and handed to the study coordinator. The sample was walked back to the laboratory and processed by standard methods. Plasma was immediately divided into small aliquots and stored at −80°C until the time of testing. Patient sex and race were captured as reported in the electronic medical record. Healthy controls were recruited by posted flier (HUM00044257) and information on race was not available.

### Cell culture

Human umbilical vein endothelial cells (HUVEC) were purchased from ATCC. For most experiments, HUVEC were cultured in Endothelial Cell Growth Basal Medium-2 (CC-3156, Lonza) supplemented with EGM-2 MV SingleQuots Kit (CC-4147, Lonza) without hydrocortisone. 10,000 cells/well were seeded into 0.2% gelatin-coated 96-well tissue culture plates the day before experiments. The following day, HUVEC were cultured with 2.5% human serum or IgG at a concentration of 100 μg/ml at 37°C in a humidified atmosphere of 5% CO_2_.

For the 2.5% human plasma experiments, HUVEC were cultured in Endothelial Cell Basal Medium (CC-3121, Lonza) supplemented with EGM-MV SingleQuots Kit (CC-4143, Lonza). All experiments were performed using HUVEC of passage 6 or fewer.

### Neutrophil adhesion assay

1×10^4^ HUVEC per well were seeded into a 96-well plate and cultured until confluent. In the BSL-3 facility, the HUVEC monolayer was cultured with 2.5% COVID serum or control serum for 6 hours. Fresh neutrophils isolated from healthy controls were labeled with calcein-AM (C1430, Thermo) for 30 minutes at 37°C. After washing the HUVEC monolayer, 3×10^5^ labeled neutrophils were then added and incubated for 20 minutes. After washing with pre-warmed HBSS, adherent neutrophil fluorescence was measured with a microplate fluorescence reader (BioTek) at 485 and 535 nm. After reading, the cells were fixed with 4% PFA for 30 minutes, and images were captured with a Cytation 5 Cell Imaging Multi-Mode Reader (BioTek) through the GFP channel.

### IgG depletion from serum

IgG was purified from COVID-19 and control serum with protein G Agarose beads (Pierce). Briefly, 800 μl of protein G agarose beads were washed at least 10 times by PBS. Then, 200 μl of serum was added and incubated with the beads at 4°C overnight. The supernatant was collected after spinning the tube at 2500xg for 5 minutes. The supernatant was the IgG depleted serum. The same volume of serum was incubated without beads under the same conditions as a mock control. Depletion was confirmed by Coomassie-stained gels.

### Purification of human IgG fractions from serum

Serum was diluted in IgG binding buffer and passed through a Protein G Agarose column (Pierce) at least 5 times. IgG was then eluted with 0.1 M glycine and neutralized with 1 M Tris. This was followed by overnight dialysis against PBS at 4°C. IgG purity was verified with Coomassie staining, and concentrations were determined by BCA protein assay (Pierce) according to manufacturer’s instructions. In some experiments, IgG was used to supplement control serum at a final concentration of 100 µg/ml. All IgG samples were determined to have endotoxin level below 0.1 EU/ml by the Pierce LAL Chromogenic Endotoxin Quantitation Kit (A39552) according to manufacturer’s instructions. This kit offers high sensitivity with linear detection range of 0.01-1.0 EU/mL.

### In-cell ELISA

In the Biosafety Level 3 facility, endothelial cell activation was assessed by an in-cell ELISA, which measured surface expression of E-selectin, ICAM-1, VCAM-1 on endothelial cells. Briefly, confluent monolayers of HUVEC in 96-well microplates were incubated with 2.5% serum or 100 µg/ml purified IgG for 6 hours and then fixed using the same volume of 8% paraformaldehyde for 30 minutes. Cells were blocked with 2x blocking solution (Abcam) at room temperature for 2 hours. After washing with PBS, cells were incubated with 5 µg/ml primary mouse anti-human antibodies against E-selectin (catalog BBA26, R&D), VCAM-1 (catalog BBA5, R&D), or ICAM-1 (ab2213, Abcam) at 4°C overnight. Next, 100 µl of diluted horseradish peroxidase conjugated rabbit anti-mouse IgG (1:2000, Jackson ImmunoResearch) in 1x blocking solution was added and incubated at room temperature for 1 hour. After washing thoroughly, 100 µl of TMB substrate was added blue color development was measured at OD 650 nm with a Cytation 5 Cell Imaging Multi-Mode Reader (BioTek). The signals were corrected by subtracting the mean signal of wells incubated in the absence of the primary antibody from all other readings. Fold change was calculated by normalizing experimental wells to the mean signal of wells incubated with culture medium alone.

### Quantification of soluble E-selectin and soluble ICAM-1 in serum

Soluble E-selectin and ICAM-1 were quantified using E-selectin ELISA (DY724, R&D Systems) and ICAM-1 ELISA (DY720, R&D Systems) kits according to the manufacturer’s instructions.

### Quantification of antiphospholipid antibodies (aPL)

aPL were quantified in serum or plasma using Quanta Lite® ACA IgG, ACA IgM, aβ_2_GPI IgG, aβ_2_GPI IgM, aPS/PT IgG, and aPS/PT IgM kits (Inova Diagnostics Inc.) according to the manufacturer’s instructions, as we did previously (1). All assays are approved for clinical use and received 510(k) clearance from the FDA. Quanta Lite® aPL ELISAs (Inova Diagnostics) are well recognized by the international APS research community and are utilized by the largest international APS clinical research network registry, APS ACTION, in its core laboratories as the “gold standard” for aPL testing (2, 3). Here, aCL assays were reported in GPL and MPL units; aβ_2_GPI assays in standard β_2_GPI IgG units (SGU) and standard β_2_GPI IgM units (SMU); and aPS/PT assays in IgG and IgM units. All were per the manufacturer’s specifications.

### Quantification of myeloperoxidase-DNA complexes

Myeloperoxidase-DNA complexes were quantified similarly to what has been previously described (4). This protocol used several reagents from the Cell Death Detection ELISA kit (Roche). First, a high-binding EIA/RIA 96-well plate (Costar) was coated overnight at 4°C with anti-human myeloperoxidase antibody (Bio-Rad 0400-0002), diluted to a concentration of 1 µg/ml in coating buffer (Cell Death kit). The plate was washed two times with wash buffer (0.05% Tween 20 in PBS), and then blocked with 4% bovine serum albumin in PBS (supplemented with 0.05% Tween 20) for 2 hours at room temperature. The plate was again washed five times, before incubating for 90 minutes at room temperature with 10% serum or plasma in the aforementioned blocking buffer (without Tween 20). The plate was washed five times, and then incubated for 90 minutes at room temperature with 10x anti-DNA antibody (HRP-conjugated; from the Cell Death kit) diluted 1:100 in blocking buffer. After five more washes, the plate was developed with 3,3’,5,5’-Tetramethylbenzidine (TMB) substrate (Invitrogen) followed by a 2N sulfuric acid stop solution. Absorbance was measured at a wavelength of 450 nm using a Cytation 5 Cell Imaging Multi-Mode Reader (BioTek). Data were normalized to *in vitro*-prepared NET standards included on every plate, which were quantified based on their DNA content.

### Quantification of cell-free DNA

Cell-free DNA was quantified in serum using the Quant-iT PicoGreen dsDNA Assay Kit (Invitrogen, P11496) according to the manufacturer’s instructions.

### Quantification of Cit-H3

Cit-H3 was quantified in serum using the Citrullinated Histone H3 (clone 11D3) ELISA Kit (Cayman, 501620) according to the manufacturer’s instructions.

### Quantification of S100A8/A9 (calprotectin)

Calprotectin levels were measured with the Human S100A8/S100A9 Heterodimer DuoSet ELISA (DY8226-05, R&D Systems) according to the manufacturer’s instructions, as we have done previously (5).

## Statistical analysis

Normally-distributed data were analyzed by 2-sided t test and skewed data were analyzed by Mann-Whitney test when two groups were analyzed. For more than two groups, one-way ANOVA or Kruskal-Wallis test with correction for multiple comparisons was utilized. Correlations were tested by Spearman’s correlation coefficient. These analyses were with GraphPad Prism software version 8, and statistical significance was defined as p<0.05. Univariate and multivariate logistic regression analysis was used to identify potential relationships between anticardiolipin IgG/M, anti-β_2_GPI IgG/M, anti-PS/PT IgG/M and HUVEC surface activation markers. Odds ratio (OR) and 95% confidence intervals (95% CI) were calculated. All variables were included in the regression model. Variables with p <0.1 by univariate analysis were entered in multivariate analysis. A 2-sided p-value <0.05 was considered statistically significant. This analysis was performed using SPSS version 23.0 (IBM, Armonk, NY, USA).

---

**Supplemental Table 1:** Patient characteristics (serum)

| **Supplemental Table 1: Patient characteristics (serum)** |  |  |  |
| --- | --- | --- | --- |
|  | **COVID-19**<br>(primary cohort) | **COVID-19**<br>(expansion cohort for<br>surface ICAM-1<br>measurement) | **Control** |
| **Demographics** |  |  |  |
| Number | 118 | 126 | 40 |
| Age (years)<sup>a</sup> | 61.9 ± 17.2 (25-96) | 58.3 ± 16.7 (16-90) | 38.3 ± 12.1 (21-64) |
| Hospital day<sup>b</sup> | 4. (1-25) | 4 ± 5.8 (1-40) |  |
| Female | 55 (47%) | 50 (40%) | 29 (73%) |
| Black/African American | 50 (42%) | 52 (41%) |  |
| White | 47 (40%) | 55 (44%) |  |
| **Comorbidities** |  |  |  |
| Diabetes | 45 (38%) | 51 (40%) |  |
| Ischemic heart disease | 36 (31%) | 67 (53%) |  |
| Renal disease | 33 (28%) | 55 (44%) |  |
| Lung disease | 29 (25%) | 67 (53%) |  |
| Autoimmune | 3 (3%) | 3 (2%) |  |
| Cancer | 15 (13%) | 17 (13%) |  |
| History of stroke | 11 (9%) | 5 (4%) |  |
| Obesity | 50 (42%) | 67 (53%) |  |
| Hypertension | 73 (62%) | 63 (50%) |  |
| Immune deficiency | 7 (6%) | 35 (28%) |  |
| History of smoking | 23 (19%) | 29 (23%) |  |
| **Respiratory status** |  |  |  |
| Mechanical ventilation | 41 (35%) | 59 (47%) |  |
| High-flow oxygen | 8 (7%) | 12 (10%) |  |
| Nasal-cannula oxygen | 38 (32%) | 29 (23%) |  |
| Room air | 31 (26%) | 26 (21%) |  |
| **Key medications** |  |  |  |
| Dexamethasone | 0 (0%) | 3 (2%) |  |
| Remdesivir | 1 (1%) | 2 (2%) |  |
| <sup>a</sup> Mean ± standard deviation (range) |  |  |  |
| <sup>b</sup> Median = 3 (original) and 3 (expansion) |  |  |  |

**Supplemental Table 2:** Patient characteristics (plasma)

| **Supplemental Table 2: Patient characteristics (plasma)** |  |  |  |
| --- | --- | --- | --- |
|  | **Sepsis** | **COVID-19** | **Control** |
| **Demographics** |  |  |  |
| Number | 100 | 72 | 38 |
| Age (years)<sup>a</sup> | 62.6 ± 6.0 (50-73) | 61.43 ± 18 (25-96) | 43.2 ± 9.3 (25-62) |
| Hospital day<sup>b</sup> | 6.7 ± 6.9 (1-40) | 5.1 ± 6.1 (0-24) |  |
| Female | 47 (47%) | 33 (46%) | 22 (58%) |
| Black/African American | 4 (4%) | 32 (44%) |  |
| White | 92 (92%) | 26 (36%) |  |
| **Comorbidities** |  |  |  |
| Diabetes | 34 (34%) | 29 (40%) |  |
| Ischemic heart disease | 26 (26%) | 25 (35%) |  |
| Renal disease | 51 (51%) | 19 (26%) |  |
| Lung disease | 42 (42%) | 16 (22%) |  |
| Autoimmune | 9 (9%) | 2 (3%) |  |
| Cancer | 37 (37%) | 8 (11%) |  |
| Obesity | 50 (50%) | 32 (44%) |  |
| Hypertension | 48 (48%) | 43 (60%) |  |
| History of smoking | 49 (49%) | 15 (21%) |  |
| **Respiratory status** |  |  |  |
| Mechanical ventilation | 65 (65%) | 33 (46%) |  |
| High-flow oxygen | 11 (11%) | 5 (7%) |  |
| Nasal-cannula oxygen | 24 (24%) | 21 (29%) |  |
| Room air | 0 (0%) | 13 (18%) |  |
| <sup>a</sup> Mean ± standard deviation (range) |  |  |  |
| <sup>b</sup> Median = 4 (sepsis) and 2 (COVID-19) |  |  |  |

**Supplemental Table 3:** Prevalence of antiphospholipid antibodies in serum from patients with COVID-19 (n=118)

| aPL | Number of positive<br>(manufacturer's cutoff) | % | Number of positive<br>(titer $\geq 40$ units) | % |
| --- | --- | --- | --- | --- |
| aCL IgG | 4 | 3% | 0 | 0% |
| aCL IgM | 29 | 25% | 9 | 8% |
| a$\beta_2$GPI IgG | 2 | 2% | 2 | 2% |
| a$\beta_2$GPI IgM | 5 | 4% | 3 | 3% |
| aPS/PT IgG | 28 | 24% | 15 | 13% |
| aPS/PT IgM | 18 | 15% | 12 | 10% |
| Any positive | 53 | 45% | 30 | 25% |
| The manufacturer's cutoff: aCL IgG >20 GPL, aCL IgM >20 MPL, a$\beta_2$GPI IgG >20 SGU, a$\beta_2$GPI IgM >20 SMU, aPS/PT IgG >30 IgG units, and aPS/PT IgM >30 IgM units; aPL=antiphospholipid antibodies; aCL=anticardiolipin antibodies; a$\beta_2$GPI=anti-beta-2 glycoprotein I antibodies; aPS/PT=anti-phosphatidylserine/prothrombin antibodies. | | | | |

**Supplemental Table 4:** Correlation amongst antiphospholipid antibodies in serum from patients with COVID-19 (n=118)

| | aCL IgG | | aCL IgM | | a$\beta_2$GPI IgG | | a$\beta_2$GPI IgM | | aPS/PT IgG | | aPS/PT IgM | |
| --- | --- | --- | --- | --- | --- | --- | --- | --- | --- | --- | --- | --- |
|  | r | p | r | p | r | p | r | p | r | p | r | p |
| aCL IgG |  |  | 0.44 | **** | 0.10 | ns | -0.04 | ns | 0.51 | **** | 0.15 | ns |
| aCL IgM | 0.44 | **** |  |  | 0.11 | ns | 0.14 | ns | 0.35 | **** | 0.53 | **** |
| a$\beta_2$GPI IgG | 0.10 | ns | 0.11 | ns | | | 0.25 | ** | 0.03 | ns | 0.23 | * |
| a$\beta_2$GPI IgM | -0.04 | ns | 0.14 | ns | 0.25 | ** | | | -0.06 | ns | 0.31 | *** |
| aPS/PT IgG | 0.51 | **** | 0.35 | **** | 0.03 | ns | -0.06 | ns |  |  | 0.19 | * |
| aPS/PT IgM | 0.15 | ns | 0.53 | **** | 0.23 | * | 0.31 | *** | 0.19 | * |  |  |

aCL=anticardiolipin antibodies; a $\beta_2$ GPI=anti-beta-2 glycoprotein I antibodies; aPS/PT=anti-phosphatidylserine/prothrombin antibodies; ns=not significant; \*p<0.05, \*\*p<0.01, \*\*\*p<0.001, and \*\*\*\*p<0.0001 by Spearman's method.

**Supplemental Table 5:** Correlation of antiphospholipid antibodies with clinical and laboratory data (n=118)

|  | aCL IgG |  | aCL IgM |  | aβ<sub>2</sub>GPI IgG |  | aβ<sub>2</sub>GPI IgM |  | aPS/PT IgG |  | aPS/PT IgM |  |
| --- | --- | --- | --- | --- | --- | --- | --- | --- | --- | --- | --- | --- |
|  | r | p | r | p | r | p | r | p | r | p | r | p |
| FiO<sub>2</sub> | 0.20 | * | 0.05 | ns | 0.12 | ns | -0.05 | ns | 0.15 | ns | 0.12 | ns |
| D-dimer | 0.15 | ns | 0.21 | * | -0.02 | ns | 0.004 | ns | 0.13 | ns | 0.08 | ns |
| ANC | 0.23 | * | 0.21 | * | 0.01 | ns | 0.01 | ns | 0.07 | ns | 0.20 | * |
| calprotectin | 0.29 | ** | 0.24 | ** | 0.17 | ns | 0.06 | ns | 0.23 | * | 0.33 | *** |
| MPO-DNA | 0.23 | * | 0.30 | *** | 0.23 | * | 0.08 | ns | 0.13 | ns | 0.36 | **** |

aCL=anticardiolipin antibodies; aβ<sub>2</sub>GPI=anti-beta-2 glycoprotein I antibodies; aPS/PT=anti-phosphatidylserine/prothrombin antibodies; ANC=absolute neutrophil count; MPO-DNA=myeloperoxidase-DNA complexes (NET remnants); ns=not significant; \*p<0.05, \*\*p<0.01, \*\*\*p<0.001, and \*\*\*\*p<0.0001 by Spearman's method.

**Supplemental Table 6:** Prevalence of antiphospholipid antibodies in serum from healthy controls (n=38)

| aPL | Number of positive<br>(manufacturer's cutoff) | % | Number of positive<br>(titer $\geq 40$ units) | % |
| --- | --- | --- | --- | --- |
| aCL IgG | 0 | 0% | 0 | 0% |
| aCL IgM | 1 | 3% | 0 | 0% |
| a$\beta_2$GPI IgG | 0 | 0% | 0 | 0% |
| a$\beta_2$GPI IgM | 1 | 3% | 0 | 0% |
| aPS/PT IgG | 0 | 0% | 0 | 0% |
| aPS/PT IgM | 0 | 0% | 0 | 0% |
| Any positive | 2 | 5% | 0 | 0% |
| The manufacturer's cutoff: aCL IgG >20 GPL, aCL IgM >20 MPL, a$\beta_2$GPI IgG >20 SGU, a$\beta_2$GPI IgM >20 SMU, aPS/PT IgG >30 IgG units, and aPS/PT IgM >30 IgM units; aPL=antiphospholipid antibodies; aCL=anticardiolipin antibodies; a$\beta_2$GPI=anti-beta-2 glycoprotein I antibodies; aPS/PT=anti-phosphatidylserine/prothrombin antibodies. | | | | |

**Supplemental Table 7:**
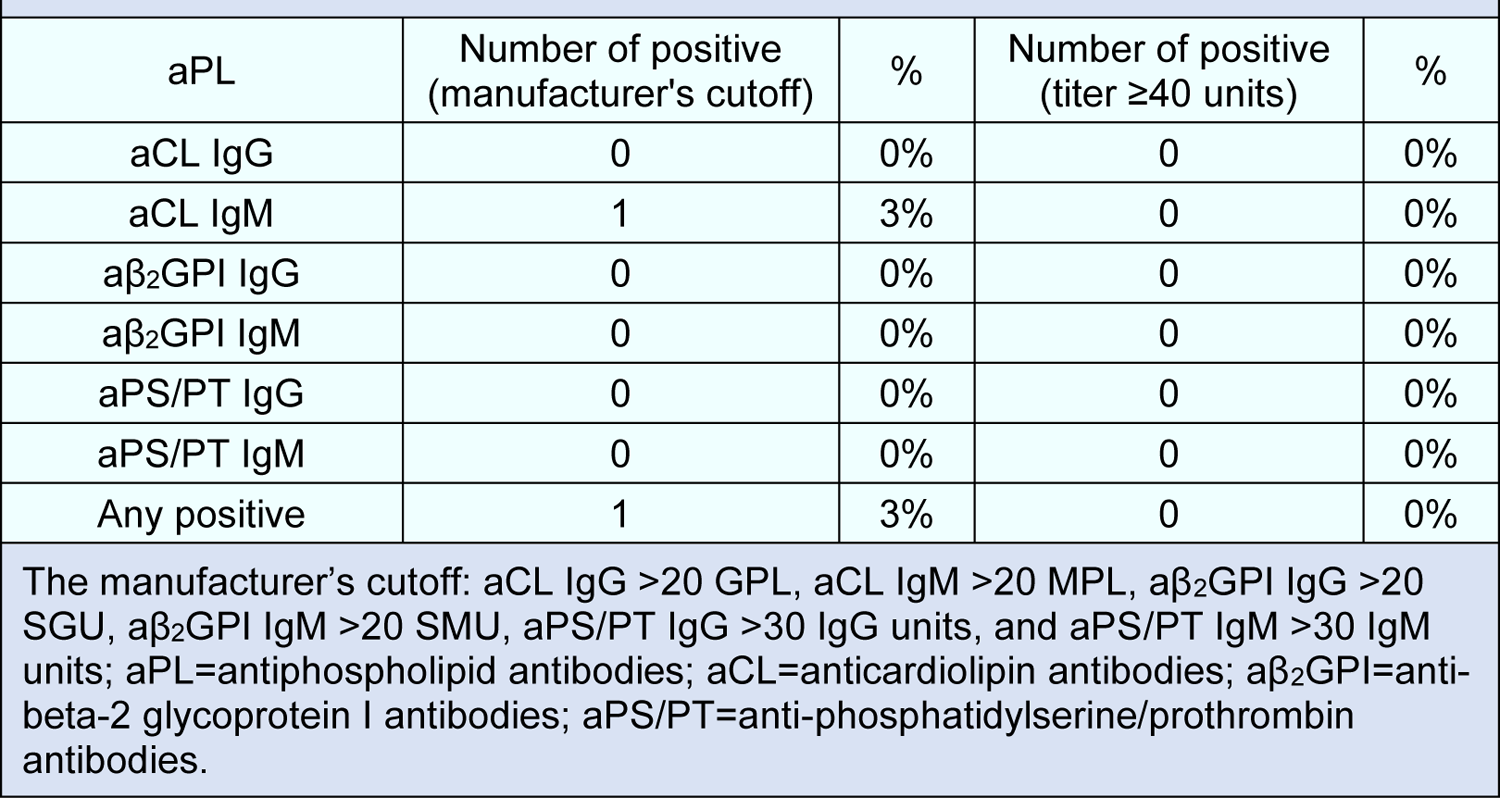
Prevalence of antiphospholipid antibodies in plasma from healthy controls (n=36)

| aPL | Number of positive<br>(manufacturer's cutoff) | % | Number of positive<br>(titer $\geq 40$ units) | % |
| --- | --- | --- | --- | --- |
| aCL IgG | 0 | 0% | 0 | 0% |
| aCL IgM | 1 | 3% | 0 | 0% |
| a$\beta_2$GPI IgG | 0 | 0% | 0 | 0% |
| a$\beta_2$GPI IgM | 0 | 0% | 0 | 0% |
| aPS/PT IgG | 0 | 0% | 0 | 0% |
| aPS/PT IgM | 0 | 0% | 0 | 0% |
| Any positive | 1 | 3% | 0 | 0% |
| The manufacturer's cutoff: aCL IgG >20 GPL, aCL IgM >20 MPL, a$\beta_2$GPI IgG >20 SGU, a$\beta_2$GPI IgM >20 SMU, aPS/PT IgG >30 IgG units, and aPS/PT IgM >30 IgM units; aPL=antiphospholipid antibodies; aCL=anticardiolipin antibodies; a$\beta_2$GPI=anti-beta-2 glycoprotein I antibodies; aPS/PT=anti-phosphatidylserine/prothrombin antibodies. | | | | |

**Supplemental Table 8:** Logistic regression analysis of adhesion molecule upregulation. Positive/negative threshold defined as 2 standard deviations above the control mean.

|  | **E-selectin** |  | univariate |  | multivariate |  |
| --- | --- | --- | --- | --- | --- | --- |
|  | HUVEC+<br>(n=52) | HUVEC-<br>(n=66) | p | OR (95% CI) | p | OR (95% CI) |
| aCL IgG (GPL) | 9.3 | 5.7 | **0.001** | 1.21 (1.08-1.35) | **0.001** | 1.21 (1.08-1.35) |
| aCL IgM MPL) | 21.7 | 18.2 | 0.11 | 1.03 (0.99-1.07) |  |  |
| aβ2GPI IgG (SGU) | 1.7 | 1.4 | 0.85 | 1.01 (0.96-1.05) |  |  |
| aβ2GPI IgM (SMU) | 2.6 | 5.3 | 0.43 | 0.99 (0.96-1.02) |  |  |
| aPS/PT IgG (units) | 24.7 | 25.8 | 0.71 | 1.00 (0.97-1.02) |  |  |
| aPS/PT IgM (units) | 26.9 | 17.9 | 0.078 | 1.01 (1.00-1.03) |  |  |
|  | **VCAM-1** |  | univariate |  | multivariate |  |
|  | HUVEC+<br>(n=47) | HUVEC-<br>(n=71) | p | OR (95% CI) | p | OR (95% CI) |
| aCL IgG (GPL) | 9.6 | 5.8 | **0.001** | 1.22 (1.10-1.36) | **0.003** | 1.18 (1.06-1.31) |
| aCL IgM MPL) | 23.8 | 17.1 | **0.008** | 1.08 (1.02-1.14) |  |  |
| aβ2GPI IgG (SGU) | 2.2 | 1.1 | 0.45 | 1.01 (0.97-1.07) |  |  |
| aβ2GPI IgM (SMU) | 3.1 | 4.7 | 0.64 | 1.00 (0.97-1.02) |  |  |
| aPS/PT IgG (units) | 29.3 | 22.6 | **0.037** | 1.03 (1.00-1.06) |  |  |
| aPS/PT IgM (units) | 23.1 | 21.1 | 0.71 | 1.00 (0.99-1.11) |  |  |
|  | **ICAM-1** |  | univariate |  | multivariate |  |
|  | HUVEC+<br>(n=28) | HUVEC-<br>(n=90) | p | OR (95% CI) | p | OR (95% CI) |
| aCL IgG (GPL) | 8.3 | 7.0 | 0.25 | 1.05 (0.97-1.13) |  |  |
| aCL IgM MPL) | 22.6 | 18.9 | 0.15 | 1.03 (0.99-1.06) |  |  |
| aβ2GPI IgG (SGU) | 1.0 | 1.7 | 0.67 | 0.98 (0.91-1.06) |  |  |
| aβ2GPI IgM (SMU) | 1.6 | 4.8 | 0.48 | 0.98 (0.92-1.04) |  |  |
| aPS/PT IgG (units) | 32.3 | 23.1 | **0.018** | 1.03 (1.01-1.06) | **0.018** | 1.03 (1.01-1.06) |
| aPS/PT IgM (units) | 25.5 | 20.8 | 0.44 | 1.01 (0.99-1.02) |  |  |

**Supplemental Table 9:** Prevalence of antiphospholipid antibodies in plasma from patients with sepsis (n=100)

| aPL | Number of positive<br>(manufacturer's cutoff) | % | Number of positive<br>(titer $\geq 40$ units) | % |
| --- | --- | --- | --- | --- |
| aCL IgG | 0 | 0% | 0 | 0% |
| aCL IgM | 10 | 10% | 1 | 1% |
| aβ2GPI IgG | 2 | 2% | 2 | 2% |
| aβ2GPI IgM | 4 | 4% | 0 | 0% |
| aPS/PT IgG | 2 | 2% | 1 | 1% |
| aPS/PT IgM | 24 | 24% | 21 | 21% |
| Any positive | 29 | 29% | 23 | 23% |
| The manufacturer's cutoff: aCL IgG >20 GPL, aCL IgM >20 MPL, aβ2GPI IgG >20 SGU, aβ2GPI IgM >20 SMU, aPS/PT IgG >30 IgG units, and aPS/PT IgM >30 IgM units; aPL=antiphospholipid antibodies; aCL=anticardiolipin antibodies; aβ2GPI=anti-beta-2 glycoprotein I antibodies; aPS/PT=anti-phosphatidylserine/prothrombin antibodies. | | | | |

**Supplementary Table 10:** Correlation of HUVEC surface ICAM-1 with antiphospholipid antibodies in patients with sepsis

|  | ICAM-1 |  |
| --- | --- | --- |
| Spearman | r | p |
| Antiphospholipid antibodies |  |  |
| IgG anticardiolipin | 0.283 | ** |
| IgM anticardiolipin | 0.191 | ns |
| IgG anti-β2GPI | 0.056 | ns |
| IgM anti-β2GPI | 0.161 | ns |
| IgG anti-PS/PT | -0.021 | ns |
| IgM anti-PS/PT | 0.293 | ** |
| **p<0.01; ns=not significant. |  |  |

**Supplemental Figure 1:**
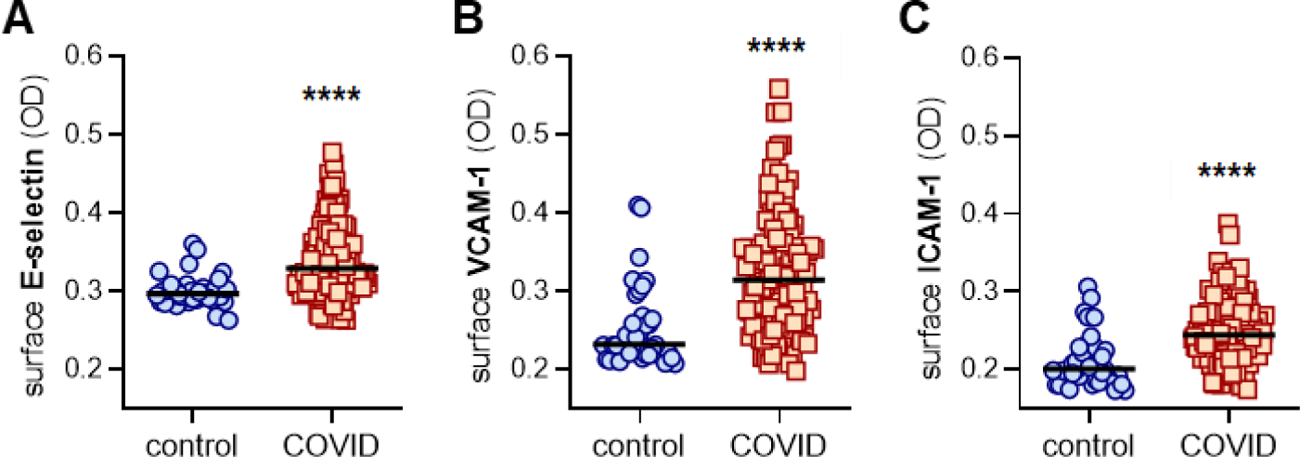
Activation of human umbilical vein endothelial cells (HUVEC) by control or COVID-19 serum (presented as optical density rather than fold change). HUVEC were cultured for 6 hours with serum from either healthy controls (collected pre-pandemic) (n=38) or patients hospitalized with COVID-19 (n=118). Cells were then fixed, and surface expression of E-selectin (**A**), VCAM-1 (**B**), or ICAM-1 (**C**) was quantified. Median values are indicated by horizontal lines. Data were presented as raw absorbance (optical density 650 nm). Groups were analyzed by Mann-Whitney test; ****p<0.0001.

**Supplemental Figure 2:**
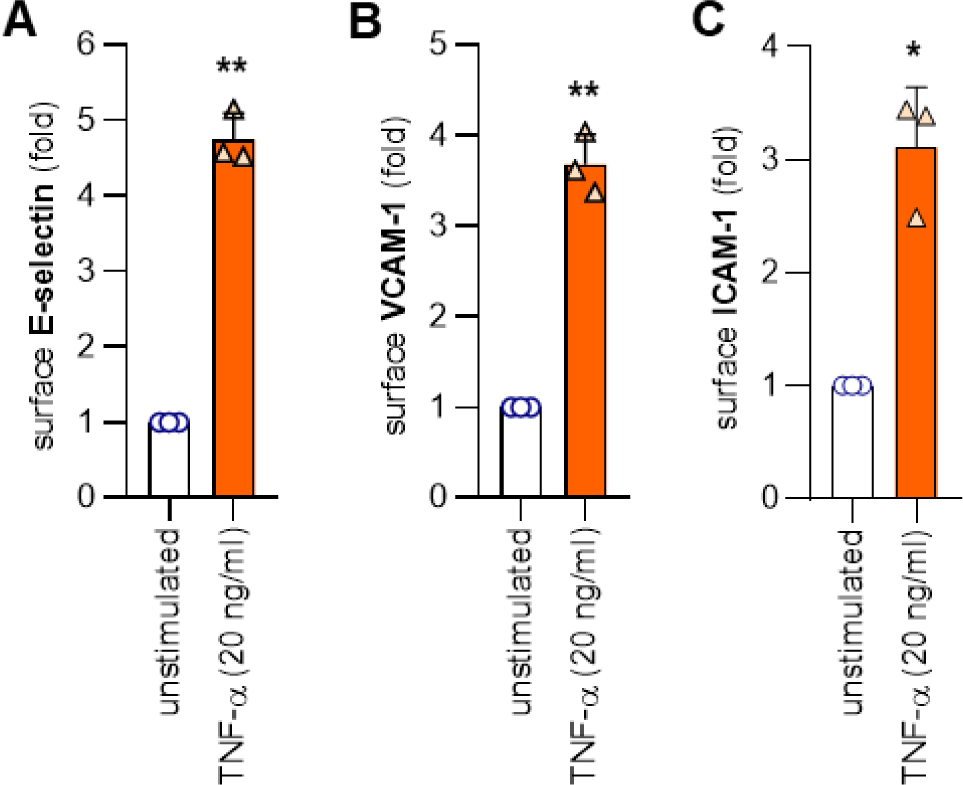
TNF-α upregulated surface adhesion molecules. **A-C**, HUVEC were cultured for 6 hours with TNF-α as indicated. Cells were then fixed, and surface expression of E-selectin (**A**), VCAM-1 (**B**), and ICAM-1 (**C**) was quantified. Mean and standard deviation are indicated for 3 independent experiments. Comparisons were by paired t test; *p<0.05 and **p<0.01.

**Supplemental Figure 3:**
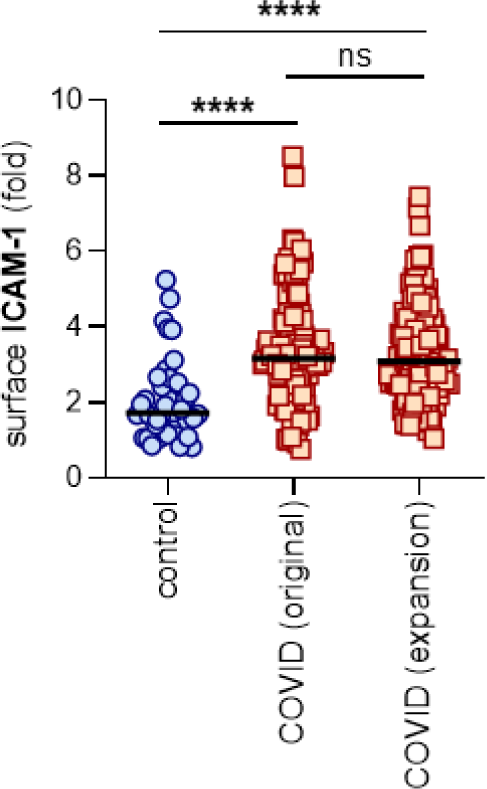
Similar ICAM-1 upregulation by original (n=118) and expansion (n=126) COVID-19 cohorts. HUVEC were cultured for 6 hours with serum from either healthy controls (collected pre-pandemic) (n=40) or patients hospitalized with COVID-19 from two similar cohorts as defined in Supplementary Table 1 (n=118 from original cohort in Figure 1D; and n=126 from expansion cohort). Cells were then fixed, and surface expression of ICAM-1 was quantified. Median values are indicated by horizontal lines. Groups were analyzed by Kruskal Wallis test with correction for multiple comparisons by Dunn’s method; ****p<0.0001 and ns=not significant.

**Supplemental Figure 4:**
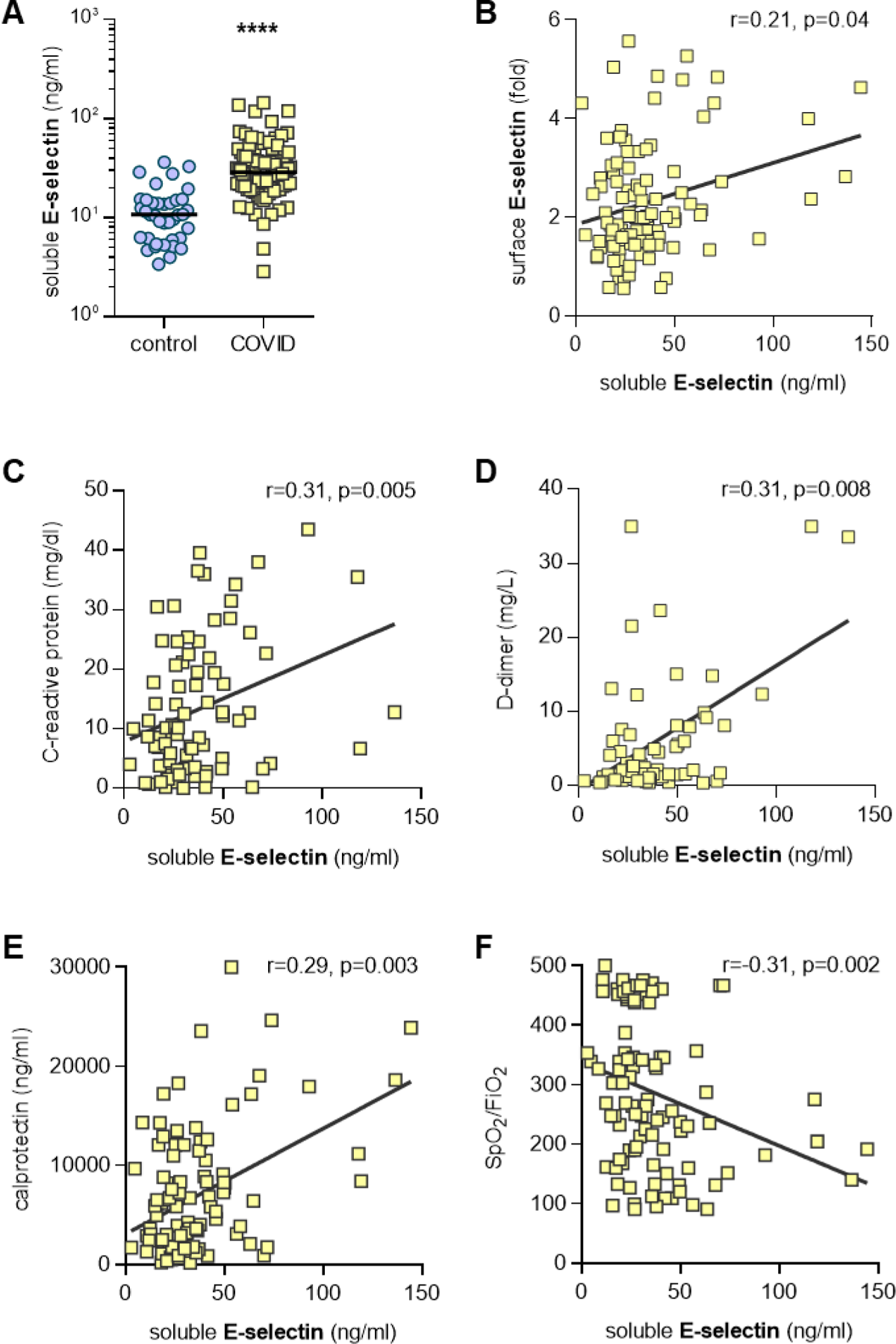
Association between soluble E-selectin in serum and clinical parameters. **A**, Serum from healthy controls (n=38) and COVID-19 patients (n=102) were assessed for soluble E-selectin. COVID-19 samples were compared to controls by Mann-Whitney test; ****p<0.0001. **B,** Soluble E-selectin was compared to HUVEC E-selectin expression as presented in Figure 1B. Correlation was determined by Spearman’s method. **C-F,** Soluble E-selectin in COVID-19 serum was compared to laboratory and clinical data when available on the same day as the serum collection. Spearman’s correlations are presented for C-reactive protein (n=83) (**C**), D-dimer (n=72) (**D**), calprotectin (n=102) (**E**), oxygenation efficiency (n=99) (pulse oximetry/fraction of inspired oxygen, **F**).

**Supplemental Figure 5:**
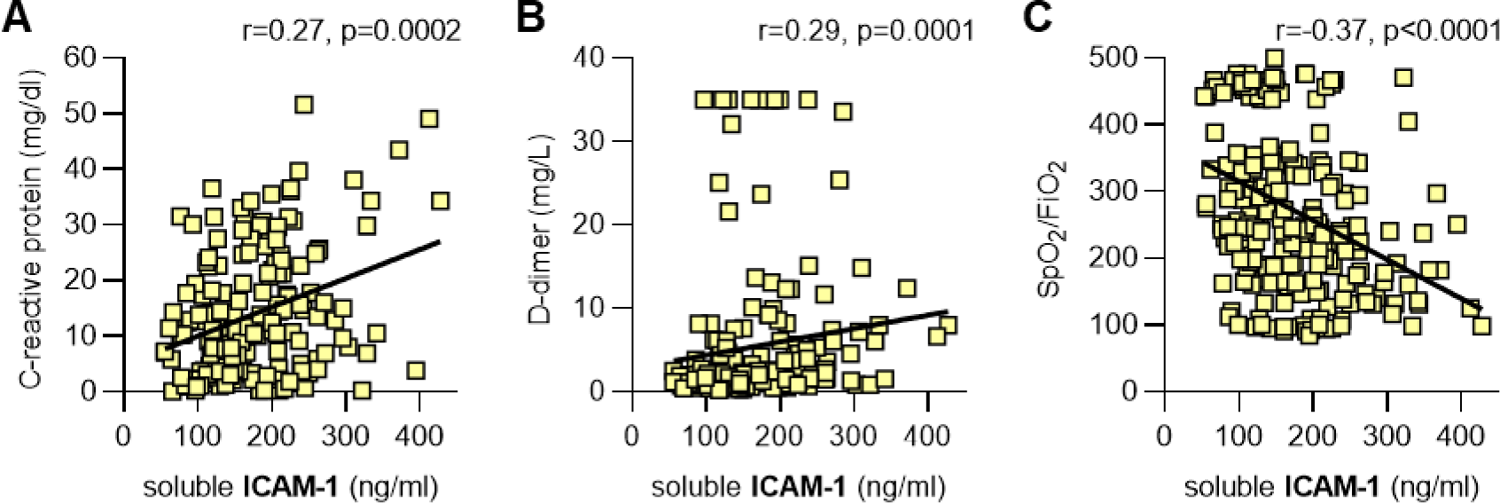
Association between soluble ICAM-1 and clinical parameters. **A-C**, Soluble ICAM-1 in COVID-19 serum was compared to laboratory and clinical data when available on the same day as the serum collection. Spearman’s correlations are presented for C-reactive protein (n=176) (**A**), D-dimer (n=174) (**B**), and oxygenation efficiency (n=228) (pulse oximetry/fraction of inspired oxygen, **C**).

**Supplemental Figure 6:**
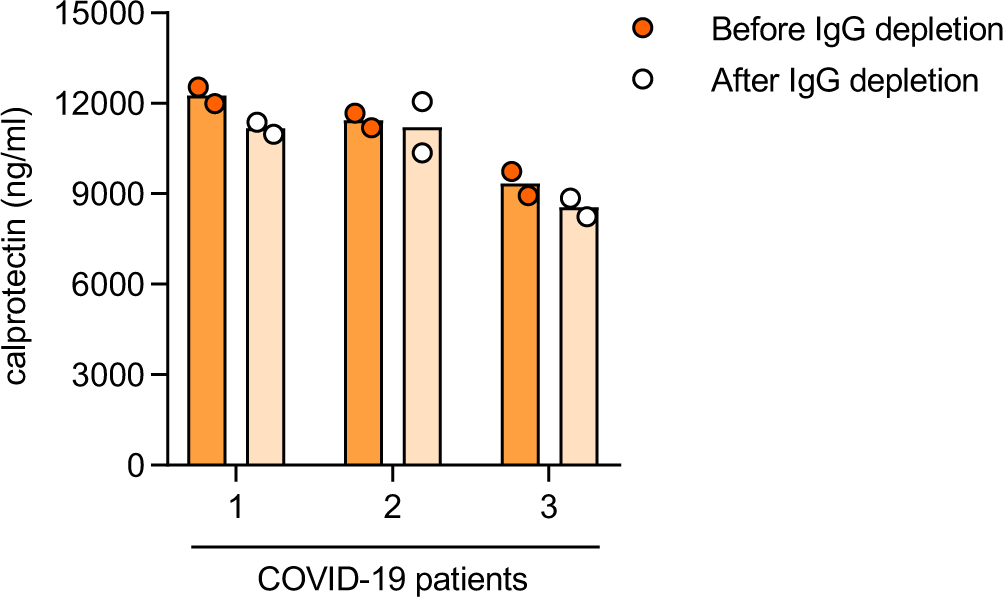
Serum calprotectin levels remain stable after IgG depletion. Serum calprotectin levels from 3 COVID-19 patients were tested before and after IgG depletion. There were no statistically significant comparisons by paired t-test.

**Supplemental Figure 7:**
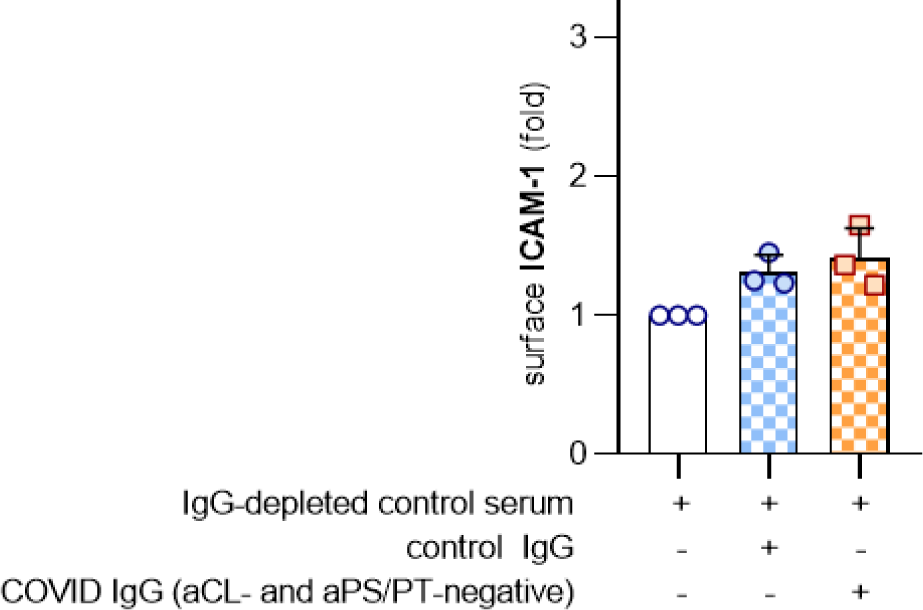
IgG from serum with negative anticardiolipin (aCL) and anti-phosphatidlyserine/prothrombin (aPS/PT) antibodies does not trigger HUVEC activation. Serum was pooled from 3 patients with negative testing for both aCL IgG and aPS/PT IgG. IgG (100 μg/ml) was purified from the pooled samples, and then spiked into control serum that had been depleted of IgG. Activation of HUVEC was determined after culture for 6 hours as defined by surface expression of ICAM-1. Groups were compared by one-way ANOVA with correction for multiple comparisons by Dunnett’s test; no comparisons were statistically significant.

